# Interpretable Trajectory-Based Feature Extraction from Longitudinal Electronic Health Records

**DOI:** 10.64898/2026.09.14.26363036

**Authors:** Vitaly Bulgakov, Alexander Turchin

## Abstract

Longitudinal electronic health records (EHRs) contain rich information about the evolution of a patient’s clinical state, but temporal measurements and events are often represented using summary statistics or complex learned representations that can be difficult to interpret clinically. We propose an interpretable trajectory-based feature representation that transforms heterogeneous longitudinal EHR data into compact, human-readable predictive features. Numerical temporal variables are represented by trajectory states such as INCREASING, DECREASING, and `STABLE`, while categorical clinical events are represented by interpretable states describing repeated occurrence, stability, or change. These trajectory features are combined with static patient and encounter characteristics and evaluated using L1-regularized logistic regression, which provides sparse feature selection and direct identification of positive and negative predictive features.

The approach is evaluated using MIMIC-IV in two clinically distinct prediction tasks: in-hospital mortality following ICU admission and hospital readmission within 30 days after discharge. For ICU mortality, multiple observation windows ranging from 1 to 48 hours are investigated to examine how the availability of longitudinal information affects prediction and the relative contribution of trajectory features. The highest AUROC of 0.8751 is obtained with a 32-hour observation window. For 30-day readmission, trajectories from the final 72 hours of the index hospitalization are evaluated using both three-state and five-state categorical representations. The two representations produce essentially identical discriminative performance (AUROC 0.6875 and 0.6876, respectively), indicating that increasing trajectory granularity provides little predictive benefit in this setting.

The results demonstrate that heterogeneous longitudinal EHR observations can be transformed into compact and directly interpretable temporal features while retaining useful predictive information. The proposed framework therefore provides a simple and model-compatible approach for identifying clinically interpretable patterns of patient evolution and can be extended with more detailed trajectory definitions or applied with other predictive models.

## 1 Introduction

We propose an interpretable trajectory-based representation for extracting clinically meaningful predictive features from longitudinal electronic health record data and evaluate it in two clinically important settings: in-hospital mortality following ICU admission and 30-day hospital readmission. Rather than focusing primarily on maximizing predictive performance, our objective is to identify compact, interpretable patterns of patient evolution that are associated with subsequent clinical outcomes and can be readily understood and potentially used by clinicians for risk assessment and decision support.

In this study, we use data from the Medical Information Mart for Intensive Care IV (MIMIC-IV) database [1], a large, publicly available, deidentified EHR dataset containing clinical information for patients admitted to Beth Israel Deaconess Medical Center (BIDMC) in Boston, Massachusetts. MIMIC-IV provides detailed data spanning both hospital admissions and intensive care stays, including patient demographics, laboratory measurements, vital signs and other charted observations, medication administrations, procedures, diagnoses, and admission, discharge, and transfer information. This combination of hospital-wide and ICU-specific longitudinal data enables the evaluation of the proposed trajectory-based representation in both study settings considered here: in-hospital mortality following ICU admission and 30-day hospital readmission.

For ICU mortality, structured clinical data collected during the first hours, e.g. 24 hours, of the ICU stay are used as the observation window. The outcome is defined as in-hospital mortality occurring at any subsequent time during the same hospitalization, including mortality after the initial observation period and, when applicable, after transfer from the ICU to another hospital unit. The resulting features therefore characterize early clinical trajectories that are predictive of mortality during the remainder of the hospitalization.

For 30-day hospital readmission, structured information from the index hospitalization is used to characterize the patient’s clinical course, while the outcome is defined as a subsequent hospital admission occurring within 30 days after discharge. This formulation separates the clinical observation period from the subsequent outcome period and allows the method to identify patterns during the index hospitalization that are associated with short-term readmission risk. The present study focuses exclusively on structured EHR data, combining structured temporal measurements and clinical events with static patient and encounter characteristics; unstructured modalities such as clinical notes and medical imaging are not considered.

Rather than representing temporal information using complete time series or summary statistics alone, numerical variables are converted into clinically meaningful trajectory categories, such as increasing, decreasing, or stable, while categorical temporal information—including medications, procedures, clinical events, and charted observations—is represented by progression states such as occurred once, occurred multiple times, changed, or remained stable. These trajectory features are combined with static patient and encounter characteristics and evaluated using sparse logistic regression.

Importantly, the predictive model serves primarily as a mechanism for identifying and validating outcome-associated features rather than as the final clinical objective itself. The resulting features directly express interpretable clinical patterns—for example, whether a laboratory measurement increased or decreased, whether a medication was administered repeatedly, or whether a clinical state changed during the observation period. Consequently, the output of the framework is not only a risk estimate but a set of human-interpretable predictive features that can be directly examined, communicated, and potentially incorporated into straightforward clinical risk assessment and decision-support workflows. The proposed approach therefore emphasizes predictive feature discovery and clinical interpretability, while retaining competitive predictive performance.

## 2 Related Work

Numerous studies have investigated mortality and readmission prediction and interpretability using structured and longitudinal electronic health record (EHR) data from MIMIC-IV and other clinical datasets. These studies employ different methodological approaches to address the temporal nature of the hospitalization process, differing substantially in how they represent the evolution of a patient’s clinical state over time and in the extent to which the resulting predictors are interpretable. In this section, we consider several of the most recent and relevant studies.

Nguyen and Mittal [2] investigated ICU mortality and length-of-stay prediction for patients with atrial fibrillation using MIMIC-IV for model development and MIMIC-III for external validation. Multiple machine learning algorithms were compared, with XGBoost providing the strongest mortality discrimination. The study primarily represents measurements collected during the ICU stay through conventional aggregated clinical features rather than explicitly encoding the direction of temporal evolution. Thus, the work focuses mainly on predictive modeling and validation rather than on constructing interpretable temporal trajectory features.

Mamandipoor et al. [3] developed a multimodal deep-learning model for predicting in-hospital mortality after the initial 24 hours of ICU admission. The approach combines structured static and time-varying clinical information with unstructured clinical notes and chest radiographs and performs extensive external validation across multiple ICU datasets. Temporal information is represented through learned model representations rather than explicit clinically interpretable trajectory categories. This contrasts with approaches that transform temporal measurements into predefined states describing the direction or pattern of clinical change.

Mamatov and Kellmeyer [4] studied in-hospital mortality prediction specifically in cardiac-arrest ICU patients using MIMIC-IV. Structured clinical measurements from the early ICU period are combined with textual information, and feature-selection methods including LASSO and XGBoost are used to identify informative predictors. Temporal measurements are largely represented through conventional summary statistics, such as minimum, maximum, and mean values, rather than through explicit trajectory states. Consequently, the study emphasizes predictive performance and feature selection rather than representation of the temporal direction of clinical variables.

Sadanandan [5] considered early prediction of patient deterioration in the ICU using MIMIC-IV. The outcome includes impending adverse events such as mortality, vasopressor initiation, and mechanical ventilation. Structured longitudinal vital signs and laboratory measurements are processed as time series and combined with representations extracted from clinical notes. A deep sequential model therefore learns temporal patterns implicitly from the measurements. This provides an important alternative to explicit trajectory engineering: temporal evolution is retained, but its representation is learned by a complex model rather than expressed directly through human-readable states such as increasing, decreasing, or stable.

Pang et al. [6] developed ICU mortality prediction models from MIMIC-IV using physiological components of the APS III and LODS severity scoring systems. Logistic regression, support vector machines, decision trees, and XGBoost were compared, with XGBoost achieving the strongest predictive performance. The approach primarily summarizes physiological severity during the ICU observation period rather than explicitly representing the evolution of individual variables over time. This work therefore demonstrates the predictive value of conventional physiological severity representations, while leaving the temporal direction of clinical change largely unmodeled.

Trajectory-based approaches have also been investigated for hospital readmission. Ben-Assuli et al. [7] provide one of the most directly relevant examples. They analyzed 30-day readmission using the developmental trajectory of creatinine as a longitudinal marker. A semi-parametric group-based trajectory model was applied to EHR data to identify three distinct creatinine trajectories associated with different readmission rates. This study demonstrates that the temporal evolution of a clinical measurement can itself provide useful readmission information. However, the trajectory analysis is centered on a single laboratory marker, whereas a general trajectory representation can potentially be applied systematically to many heterogeneous numerical and categorical EHR variables.

Jiang et al. [8] proposed a dynamic prediction framework for 30-day readmission in patients with heart failure. Rather than constructing trajectories of individual clinical variables, the model repeatedly updates readmission risk as new information becomes available during hospitalization. The resulting sequence of predicted risks is then used to characterize distinct *readmission-risk trajectories*. Thus, the trajectory is defined in the prediction space rather than directly in the underlying clinical-feature space.

Kirk et al. [9] investigated 30-day readmission after radical cystectomy using dynamic postoperative laboratory information. The authors characterized trajectories of routinely measured laboratory variables during the index hospitalization and showed that longitudinal measurements, including white blood cell count, blood urea nitrogen, bicarbonate, and creatinine, improved readmission prediction compared with models based on static information alone. This study provides evidence that changes in physiological variables during hospitalization contain predictive information that may be lost when only static measurements are considered.

Fahimi et al. [10] investigated dynamic 30-day readmission prediction for patients with heart failure using longitudinal vital signs obtained through telemonitoring. Sequential measurements such as blood pressure, heart rate, and body weight were incorporated into a recurrent neural network so that readmission risk could be updated as new observations became available. As with other sequence-learning approaches, temporal evolution is learned implicitly by the model rather than transformed into explicit, clinically interpretable trajectory categories.

Tang et al. [11] proposed a multimodal spatiotemporal graph neural network for 30-day all-cause hospital readmission prediction. The method integrates longitudinal EHR information and chest radiographs while also modeling similarity relationships among patients through a graph representation. The proposed model achieved an AUROC of approximately 0.79 on two independent datasets. This work explicitly exploits temporality, but the temporal information is encoded within a complex learned spatiotemporal representation rather than converted into directly interpretable clinical trajectory states.

Ge et al. [12] developed an interpretable ICU mortality prediction model that combines sequential clinical features, such as vital signs and laboratory measurements, with non-sequential features. The study compared logistic regression with a recurrent neural network based on long short-term memory (LSTM) units and investigated the contribution of individual clinical features to mortality prediction. Although the recurrent model captures longitudinal information directly, interpretation of the sequential features remains dependent on the learned model representation rather than on explicit human-readable descriptions of how individual clinical variables evolve over time.

In subsequent work, Ge et al. [13] used a deep-learning attention mechanism to identify temporal clinical patterns associated with ICU mortality. The approach uses attention to determine which portions of longitudinal clinical data contribute strongly to mortality prediction, providing a mechanism for identifying informative temporal patterns. This work is particularly relevant to the present study because it emphasizes the identification and interpretation of predictive temporal information. However, temporal patterns are identified through a learned attention-based model, whereas our approach explicitly transforms the evolution of individual clinical variables into predefined, directly interpretable trajectory states.

Taken together, these studies demonstrate two major strategies for exploiting temporal EHR information. One strategy summarizes longitudinal observations using static statistics or severity scores, while another retains longitudinal sequences and learns temporal representations using recurrent, multimodal, or graph-based models. Several studies additionally demonstrate the predictive value of clinical trajectories, but these are generally restricted to individual biomarkers, specific patient populations, or trajectories of the predicted risk itself.

In contrast, the trajectory representation considered in the present work systematically transforms heterogeneous temporal clinical variables into explicit and interpretable states, such as *increasing, decreasing*, and *stable* for numerical measurements, together with corresponding temporal states for categorical events. This representation is intended to preserve clinically meaningful information about patient evolution while remaining compatible with interpretable predictive models.

## 3 Method

### 3.1 Hospitalization-Related Structured Features

Both the ICU mortality and 30-day readmission analyses are based on structured clinical data generated during hospitalization. The organization, terminology, and naming conventions of these data may vary across healthcare institutions. Since the present study uses the MIMIC-IV dataset, we follow the terminology and data conventions adopted by Beth Israel Deaconess Medical Center (BIDMC). Structured clinical features are divided into two main types:

- **Static features**, which characterize the patient or hospitalization without an associated time point within the observation period;
- **Temporal (dynamic) features**, which are associated with specific times and may change during the observation period.

Both static and temporal features are further classified into two subtypes:

- **Numerical features**;
- **Categorical features**.

Static features, examples of which include patient age (numerical), gender (categorical), and discharge institution (categorical), are generally self-explanatory and, when used for outcome prediction, do not require additional temporal processing. In contrast, temporal features may have multiple values recorded during the selected observation window and therefore often require further processing to obtain a compact and interpretable representation of their evolution over time. The focus of this study is to address the complexity and ambiguity of temporal features to make them interpretable.

The feature naming in ICU and HOSP subsets of MIMIC-IV is associated with names of corresponding source tables expressed in prefixes. For example CHART means a charted clinical observation coming from the ICU chartevents table, where, e.g., CHART_22_GAUGE_DRESSING_OCCLUSIVE is a categorical temporal feature describing the documented status of the dressing associated with a 22-gauge vascular access device/catheter.

Temporal features used in the two prediction tasks are grouped into clinically meaningful categories according to their source and are identified by the corresponding feature-name prefixes, as summarized in Table 1.

**Table 1:** Temporal feature categories used in the ICU mortality and 30-day readmission tasks.

| Prediction Task | Prefix | Feature Category |
| --- | --- | --- |
| ICU mortality | CHART | Charted clinical observations |
|  | EVENT | Clinical events |
|  | LAB | Laboratory measurements |
|  | MED | Medications and administered inputs |
|  | OUTPUT | Patient outputs |
| 30-day readmission | careunit | Care-unit information |
|  | diagnoses | Diagnoses |
|  | labs | Laboratory measurements |
|  | medication | Medications |
|  | service | Hospital services |

### 3.2 Trajectory Representation of Temporal Features

For trajectory representation, a uniform observation-window duration was used across patients within each prediction task, providing a consistent basis for extracting and comparing temporal patterns. Multiple observation-window durations were explored, including, for example, the first 24 hours of the ICU stay for ICU mortality prediction and the final 72 hours of the index hospitalization for 30-day readmission prediction.

The proposed framework transforms sequences of temporal observations into compact categorical representations that describe the evolution of a clinical feature over a predefined observation window. The trajectory definitions demonstrated in this section represent a deliberately simple implementation of this general concept and are intended to demonstrate how heterogeneous longitudinal EHR data can be converted into directly interpretable features. More elaborate trajectory definitions can be incorporated into the same framework.

#### 3.2.1 Numerical Temporal Features

For a numerical temporal feature *x*, all measurements recorded within the observation window are considered. If multiple measurements are available at the same time point, they are first averaged. Let *x*_first_ and *x*_last_ denote the first and last resulting values within the observation window, respectively. The absolute change is defined as

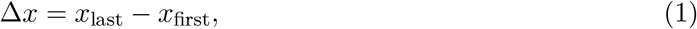

and the relative change is calculated as

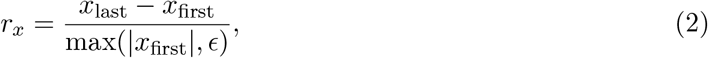

where *ϵ* is a small positive constant used to avoid division by zero.

In the simplest trajectory classifier considered here (Classifier 1), the numerical trajectory is defined as

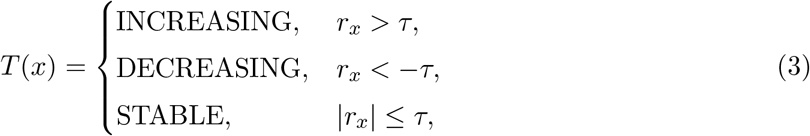

where *τ* is a predefined relative-change threshold. For example, *τ* = 0.20 corresponds to a 20% relative-change threshold.

Thus, a potentially large number of numerical measurements collected over time is represented by a single clinically interpretable trajectory category, such as INCREASING, DECREASING, or `STABLE`. This representation intentionally captures only the overall direction of change between the beginning and end of the observation window and should therefore be viewed as the simplest example of trajectory extraction rather than as an exhaustive characterization of temporal dynamics.

#### 3.2.2 Categorical Temporal Features for the Readmission case

Categorical temporal features are represented according to the type and sequence of events observed within the same observation window. Because the meaning of temporal evolution differs across clinical feature categories, simple category-specific trajectory rules are used.

For medications, the trajectory indicates whether the medication was administered once or more than once:

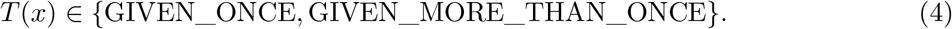

Similarly, procedures are represented as

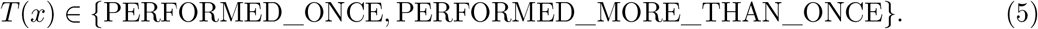

For categorical features describing patient location or hospital service, the first and last observed states are compared. The resulting trajectory is represented as `UNCHANGED` when the state remains the same and `CHANGED` when a transition occurs during the observation window.

These categorical definitions, like the numerical trajectory classifier, are intentionally simple. Their purpose is to illustrate the central idea of the proposed framework: rather than retaining a potentially complex sequence of time-stamped observations, the temporal evolution of each clinical feature is summarized by a compact and human-interpretable state that can subsequently be used directly as a predictive feature.

#### 3.2.3 Categorical Temporal Features for the ICU Mortality case

For the ICU mortality case, categorical trajectories are slightly different from those used for the 30-day readmission task, reflecting differences in the types of clinical events and observations available in the ICU data. The simple trajectory representations used for the different ICU feature types are summarized in Table 2.

**Table 2:** Simple trajectory representations for categorical temporal features in the ICU mortality task.

| ICU Feature Type | Simple Trajectory Representation |
| --- | --- |
| Medication (MED) | GIVEN_ONCE, GIVEN_MORE_THAN_ONCE |
| Procedure (PROC) | PERFORMED_ONCE, PERFORMED_MORE_THAN_ONCE |
| Clinical event (EVENT) | OCCURRED_ONCE, OCCURRED_MORE_THAN_ONCE |
| Charted observation (CHART) | STABLE, CHANGED |

For medication, procedure, and clinical-event features, the trajectory representation captures whether the corresponding event occurred once or multiple times during the observation window. For categorical charted observations, the representation indicates whether the observed state remained stable or changed during the window. These definitions constitute a simple example of categorical trajectory construction and can be extended to more detailed representations when additional aspects of temporal evolution need to be captured.

### 3.3 Feature Selection and Predictive Modeling

Following trajectory extraction, the data were transformed into a tabular representation containing one row per patient encounter and one column per predictive feature. Numerical static features were represented by their original numerical values, whereas categorical static features and categorical trajectory features were represented as categorical variables. Numerical temporal features were represented by their corresponding trajectory categories (e.g., INCREASING, DECREASING, or `STABLE`). Missing numerical trajectories were represented as `NOT_MEASURED`, while the absence of event-based categorical features was represented as `NOT_PRESENT`.

The dataset was divided into training and test sets at the patient level, ensuring that encounters belonging to the same patient could not occur in both sets. Feature eligibility was determined using the training set only, and features represented in fewer than a predefined minimum number of training encounters were excluded. The same retained feature set was then used for both training and test data.

Before model fitting, numerical features were imputed using the median and standardized to zero mean and unit variance. Categorical features were imputed using a dedicated missing-value category and transformed using one-hot encoding.

An L1-regularized logistic regression (LR) classifier was used for outcome prediction. The L1 penalty, corresponding to the Least Absolute Shrinkage and Selection Operator (LASSO), promotes sparsity by shrinking less informative feature coefficients toward zero, with some coefficients becoming exactly zero. The regularization strength was controlled by the parameter *C*, where smaller values of *C* correspond to stronger regularization. The `liblinear` solver was used to support L1-regularized logistic regression. The maximum number of optimization iterations was specified by `max_iter`. Class imbalance could additionally be accounted for using `class_weight=‘balanced’`, which assigns class weights inversely proportional to their frequencies in the training data.

After model training, the fitted LR coefficients were used to identify and rank predictive features. For a predictor *x*_*j*_ with coefficient *β*_*j*_, the logistic regression model can be expressed as

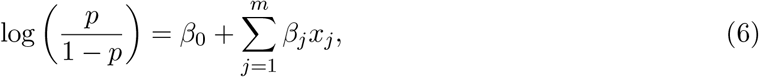

where *p* denotes the predicted probability of the outcome. Predictors with positive coefficients (*β*_*j*_ *>* 0) were identified as *positive predictive features*, indicating an association with increased probability of the outcome, whereas predictors with negative coefficients (*β*_*j*_ *<* 0) were identified as *negative predictive features*, indicating an association with decreased probability of the outcome. Within each group, features were ranked according to the magnitude of their coefficients, |*β*_*j*_|, with larger absolute coefficients indicating stronger contributions to the model prediction. Features whose coefficients were reduced to zero by L1 regularization were considered not selected by the model.

## 4 Numerical Study

The common experimental parameters are shown in Table 3.

**Table 3:** Common experimental parameters used across observation windows.

| Parameter | Value |
| --- | --- |
| Minimum encounters required for feature inclusion | 1,000 |
| Numerical trajectory relative-change threshold | 0.20 |
| Original temporal features retained | No |
| Test-set proportion | 0.20 |
| Logistic regression solver | `liblinear` |
| Regularization penalty | L1 (LASSO) |
| Inverse regularization strength ($C$) | 0.10 |

### 4.1 ICU Mortality Case

We reiterate that mortality is assessed during the remainder of the hospitalization, following the selected observation window at the beginning of the ICU stay. Experiments were conducted using the following observation window durations (in hours):

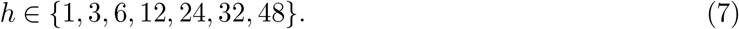

Considering multiple observation-window durations allowed us to investigate several important aspects of the proposed trajectory representation, which are discussed below. Because the duration of an ICU stay varies across patients, shorter observation windows include a larger number of eligible ICU stays, whereas longer observation windows progressively exclude stays that end before the required observation period is completed. Consequently, the 1-hour observation window contains the largest number of eligible ICU stays, see Table 4.

**Table 4:** Summary of the ICU study dataset for the smallest observation window.

| Dataset Characteristic | Count |
| --- | --- |
| Distinct ICU stays | 94,387 |
| Distinct patients | 65,328 |
| Distinct features | 2,588 |

These values decrease as the observation-window duration increases. Table 5 shows training and test set class distributions across observation-window durations. Finally, Table 6 shows the number of retained predictive features across observation-window durations.

**Table 5:** Training and test set class distributions across observation-window durations.

| Observation Window (h) | Train Class 0 | Train Class 1 | Test Class 0 | Test Class 1 |
| --- | --- | --- | --- | --- |
| 1 | 66,453 | 9,067 | 16,664 | 2,203 |
| 3 | 66,631 | 8,823 | 16,486 | 2,263 |
| 6 | 66,573 | 8,585 | 16,544 | 2,174 |
| 12 | 66,691 | 8,163 | 16,426 | 2,042 |
| 24 | 66,478 | 7,476 | 16,639 | 1,820 |
| 32 | 66,556 | 7,083 | 16,561 | 1,718 |
| 48 | 66,443 | 6,378 | 16,674 | 1,613 |

**Table 6:** Number of retained predictive features across observation-window durations.

| Observation Window (h) | Number of Features |
| --- | --- |
| 1 | 372 |
| 3 | 499 |
| 6 | 565 |
| 12 | 612 |
| 24 | 669 |
| 32 | 687 |
| 48 | 705 |

#### 4.1.1 Training the Model with Logistic Regression

Model training and test parameters are shown in Table 3. Test result metrics for every observation window are shown in Figure 1. In terms of metrics the 24 hours observation window case seems to be optimal, but the highest AUROC was achieved with 32 hours window where it is slightly higher that with 24 hours case. It is *AUROC* = 0.8751 and accuracy is 0.8875

**Figure 1:**
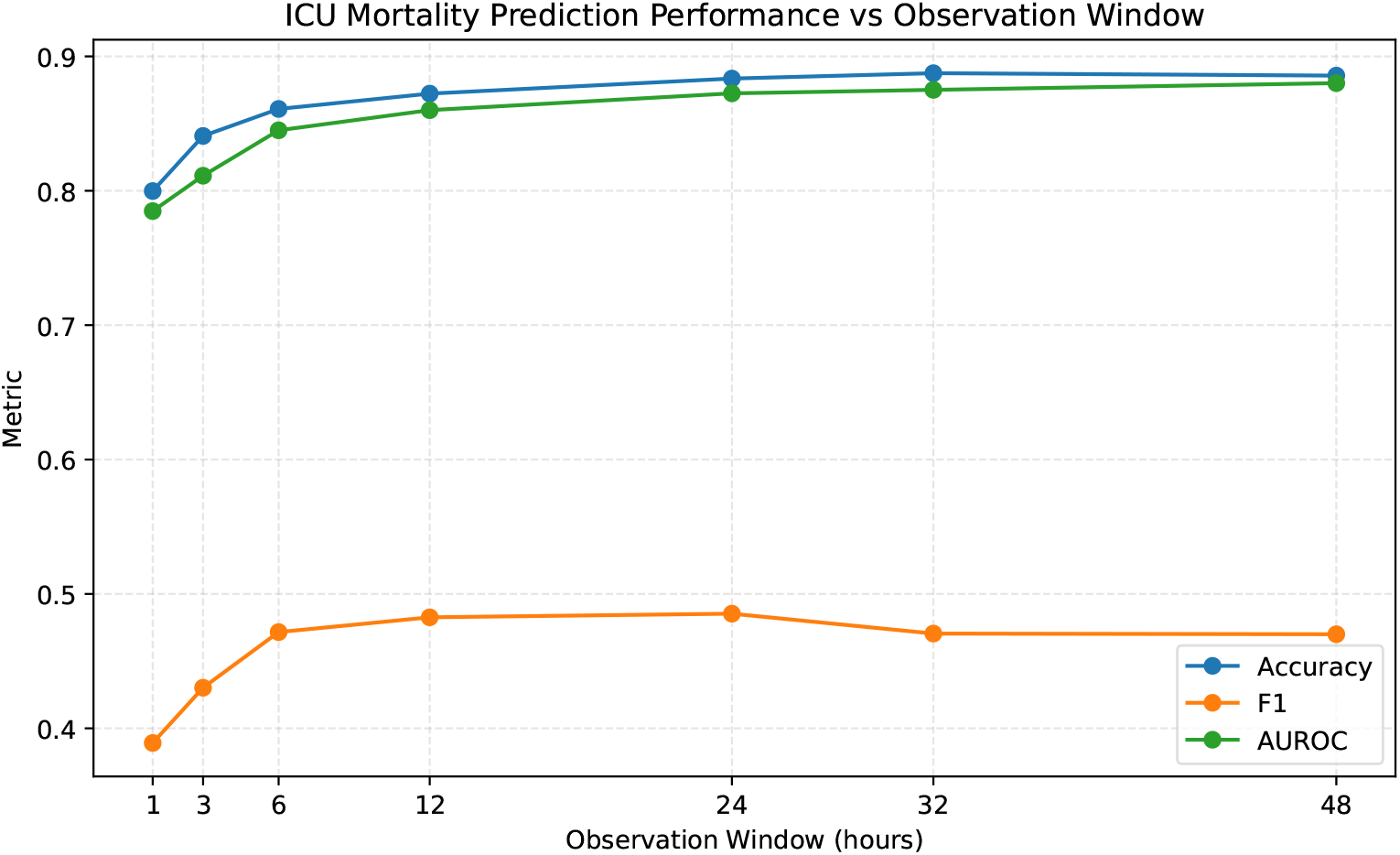
Prediction metrics vs observation window duration.

#### 4.1.2 Predictive Contribution of Static and Trajectory Features

Let *β*_*j*_(*h*) denote the logistic regression coefficient associated with predictive feature *j* for an observation window of duration *h*. After excluding temporal features represented by missing or absent states, such as `NOT_MEASURED` and `NOT_PRESENT`, the remaining predictive features are divided into two sets: T (*h*), containing observed temporal trajectory features, and S(*h*), containing static features.

The total coefficient magnitude associated with observed trajectory features is defined as

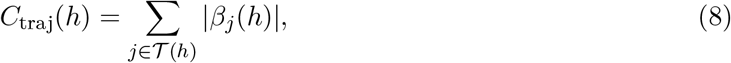

whereas the corresponding magnitude for static features is

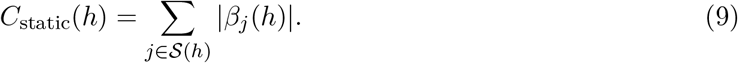

The relative contribution of observed trajectory features is then defined as

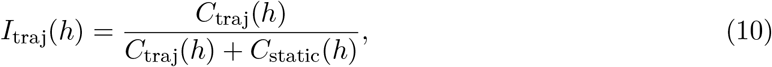

and the relative contribution of static features as

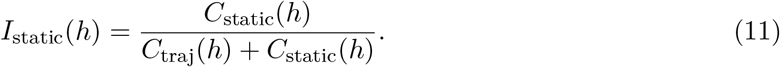

By construction,

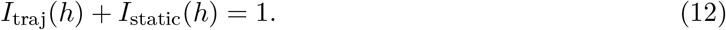

These quantities provide a simple measure of the relative contribution of temporal trajectory information and static patient information to the fitted logistic regression model at different observation-window durations. Examining *I*_traj_(*h*) as a function of *h* allows us to evaluate whether the relative predictive contribution of temporal trajectories increases as a longer period of the patient’s ICU course becomes available. The absolute coefficient values are used because both positive and negative coefficients may represent important predictive information, while their sign indicates the direction of association with the predicted outcome.

Based on these considerations, we constructed separate graphs for positive and negative predictive features, showing the dependence of *I*_traj_(*h*) and *I*_static_(*h*) on the observation-window duration *h*. These quantities characterize the relative contributions of temporal trajectory and static features to the prediction model; see Figures 2 and 3. The results demonstrate the substantial contribution of temporal trajectory features to the model and illustrate how their relative importance evolves with increasing observation-window duration.

**Figure 2:**
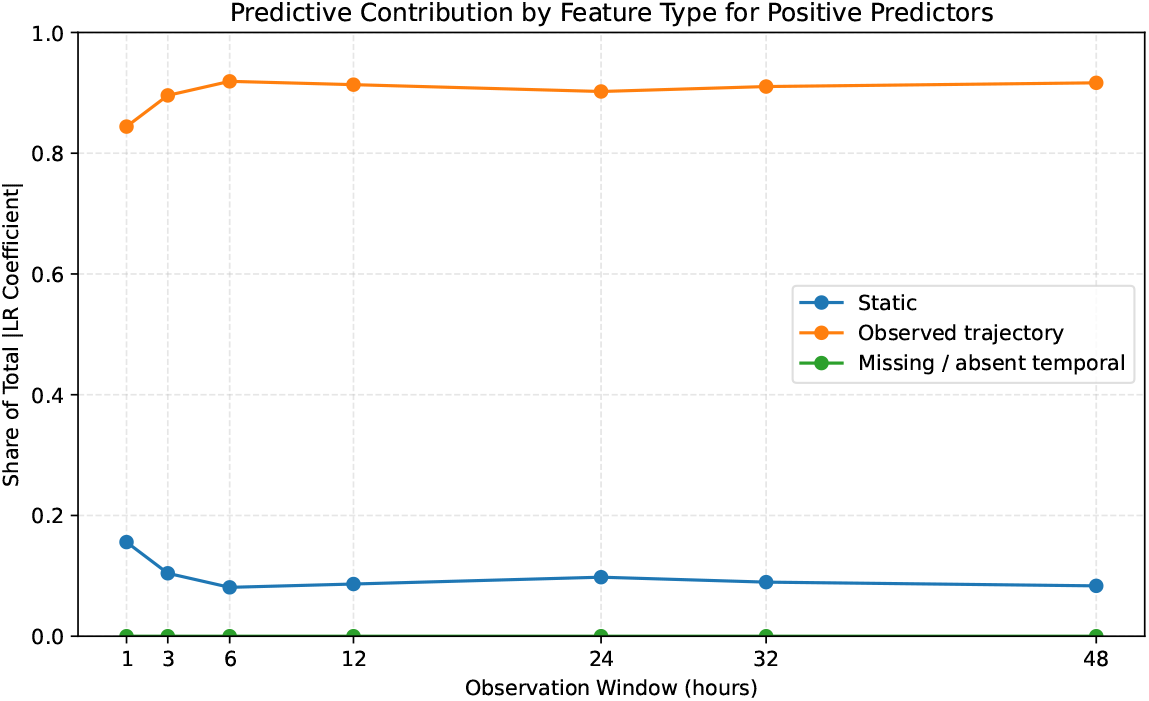
Feature types contribution to prediction vs observation window duration for positive cases.

**Figure 3:**
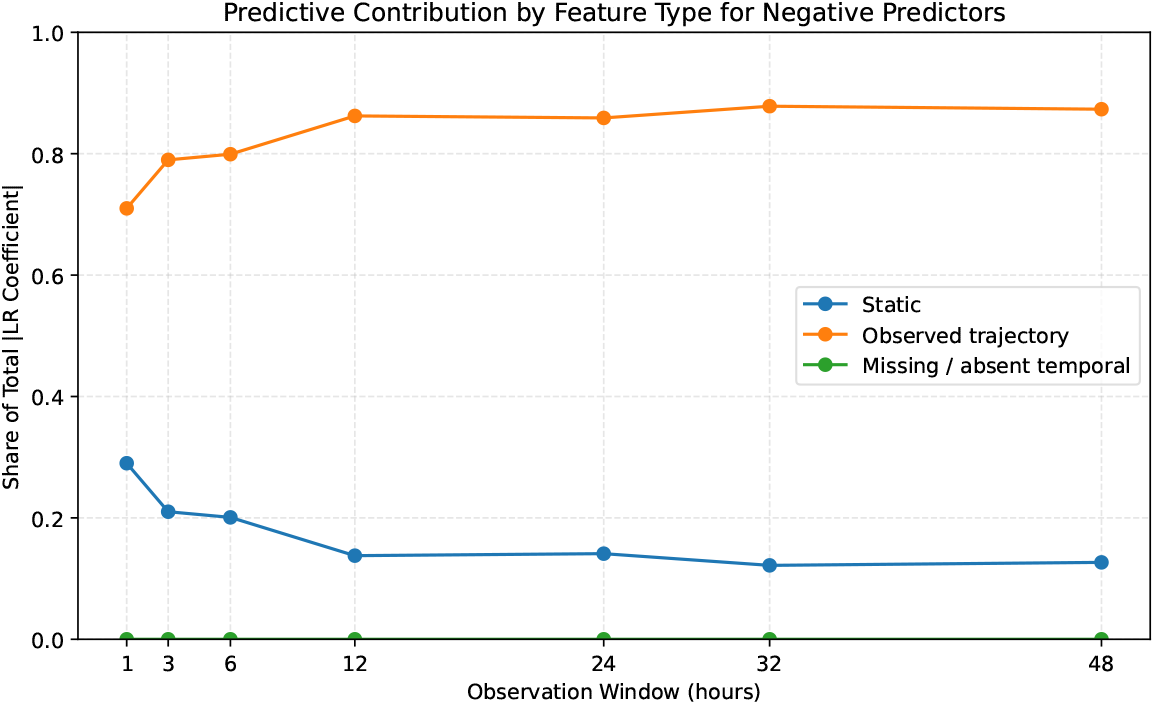
Feature types contribution to prediction vs observation window duration for negative cases.

#### 4.1.3 Selection of features with the most predictive power

Logistic regression (LR) assigns a coefficient to each predictive feature, with the coefficient magnitude reflecting its contribution to the model prediction and its sign indicating the direction of the association. *L*1 regularization (LASSO) promotes feature selection by shrinking coefficients of less informative predictors toward zero, with many becoming exactly zero, thereby retaining a sparse set of features with the strongest predictive contributions.

Figure 4 shows the top positive and negative predictive features for the 24-hour observation window, ranked in descending order according to the magnitude of their LR coefficients. The feature representations are directly interpretable and provide clinically meaningful descriptions of patient characteristics and temporal trajectories.

**Figure 4:**
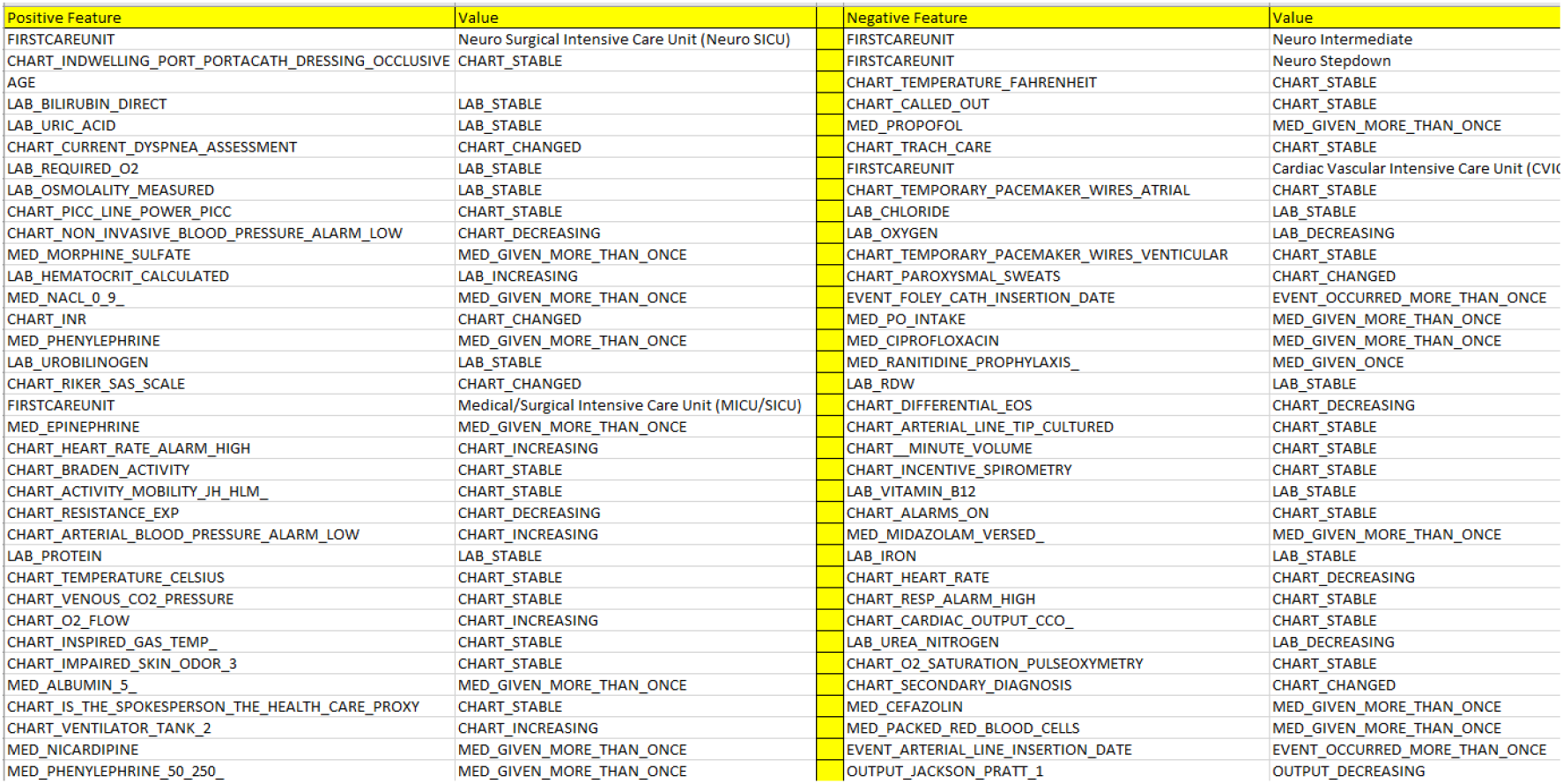
Top predictive positive and negative features for 24 hours observation window

Intuitively, one approach to identifying robust predictive features is to examine features that are consistently selected across different observation-window durations. We begin with an extreme comparison between the 1-hour and 24-hour observation windows and consider the top 15 predictive features from each. With only one hour of observation, temporal features have limited time to develop informative trajectories; therefore, we would expect the common highly ranked predictors to be dominated by static or non-trajectory information. This expectation is supported by the results: only three features are common to the top 15 predictors of both observation windows: `FIRSTCAREUNIT, AGE`, and `CHART_PICC_LINE_POWER_PICC`.

A more informative approach is to consider two or three sufficiently long observation windows, in which temporal trajectories have had adequate time to develop. Figure 5 shows the common features obtained for two observation windows (12 and 24 hours) and for three observation windows (12, 24, and 32 hours), considering the top 15 positive predictive features for each window. The comparison between the 12- and 24-hour windows identified eight common features, whereas the comparison across the 12-, 24-, and 32-hour windows identified six common features. Importantly, all six features identified in the three-window comparison were also among the eight features identified in the two-window comparison. This nested pattern indicates consistency of feature selection across observation-window durations and suggests that these features represent particularly stable predictors.

**Figure 5:**
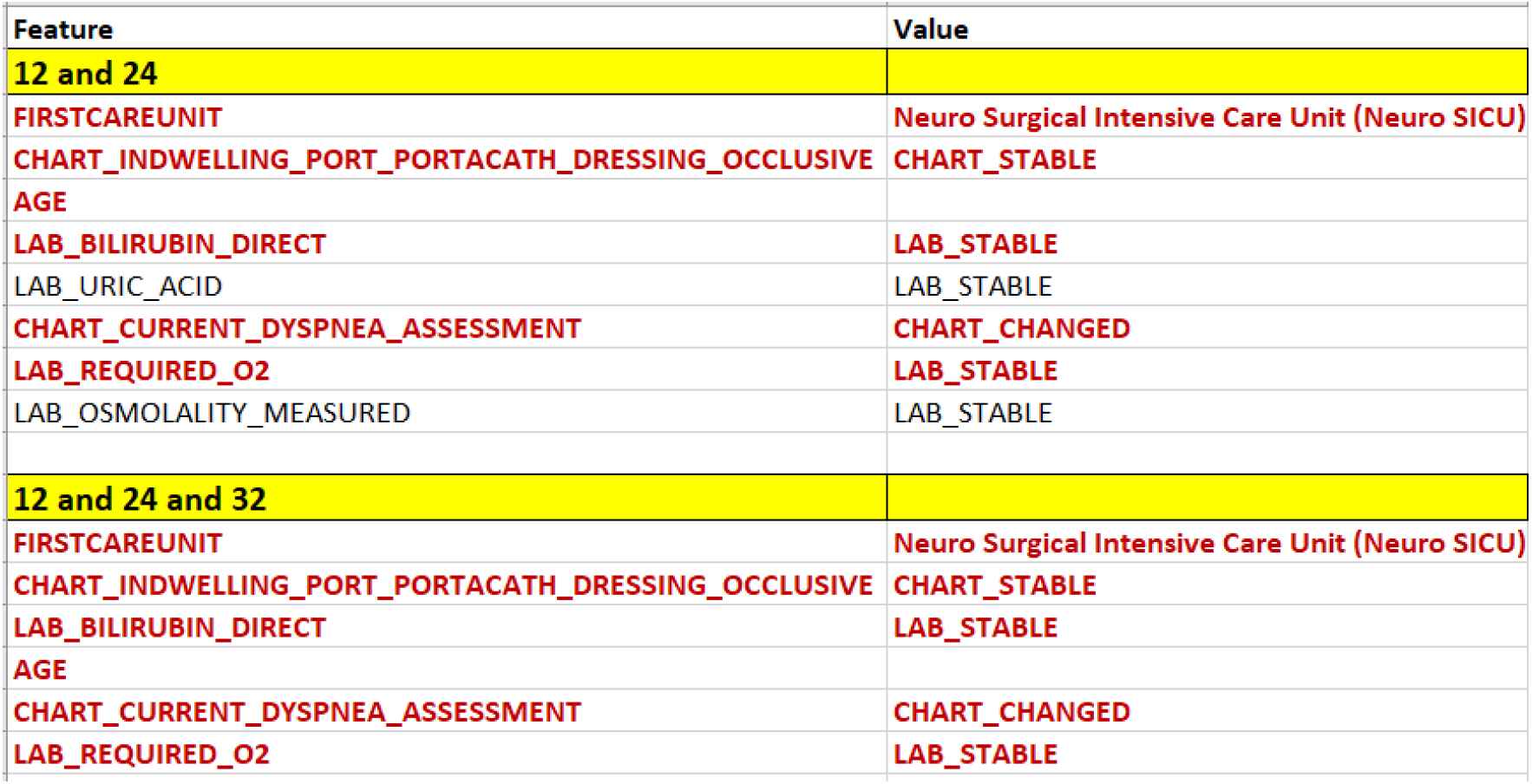
Common predictive positive features out of 15 marked RED

We conclude this section with another useful approach for enhancing the interpretability of temporal features: a graphical representation of their behavior over time, as demonstrated in Figure 6.

**Figure 6:**
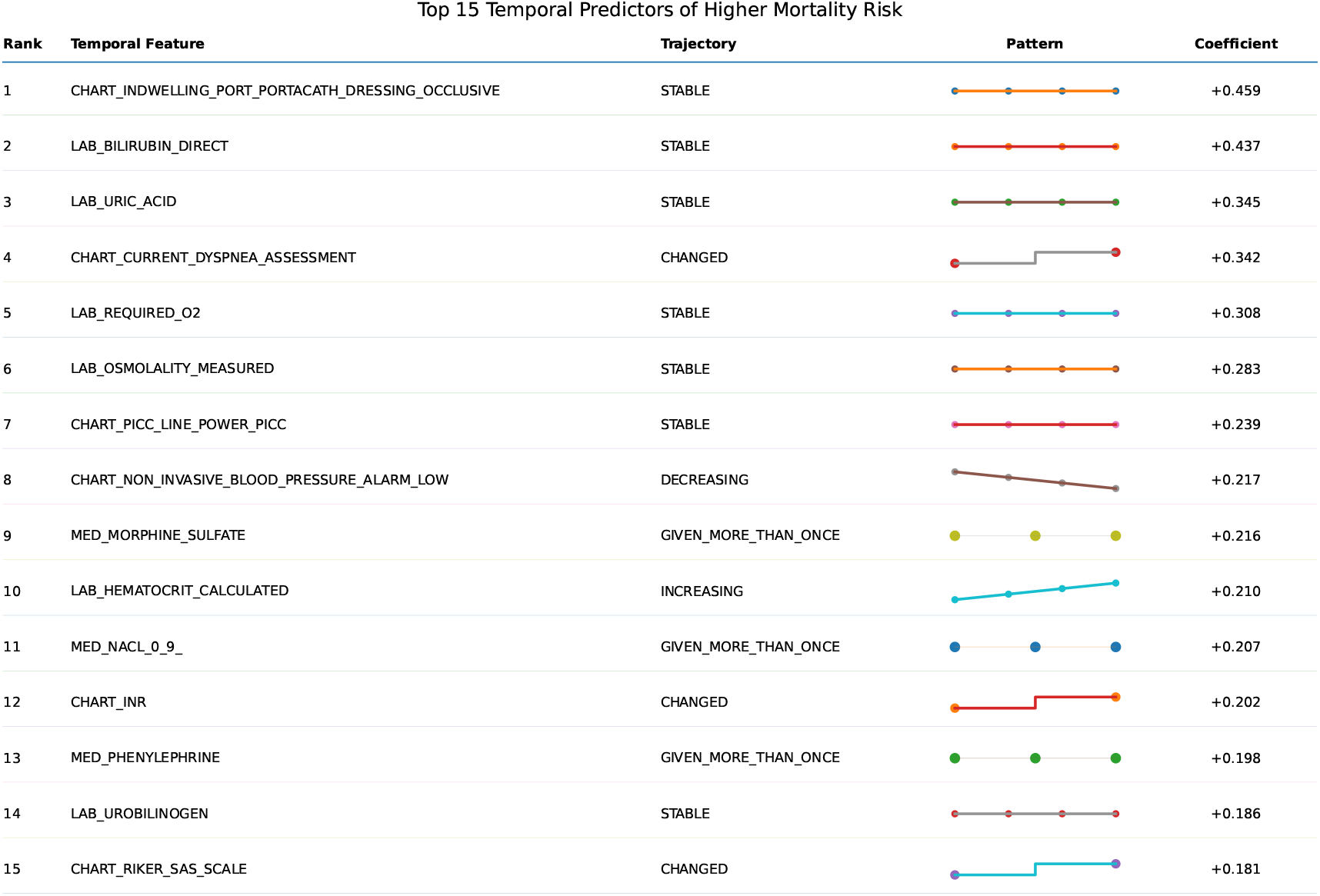
Trajectory Graphical representation of top 15 temporal positive features for 24 hours observation window

### 4.2 Readmission within 30 Days after Discharge

The second experimental case evaluates the proposed trajectory-based representation for predicting hospital readmission within 30 days after discharge. In contrast to the ICU mortality task, where the observation window is defined from the beginning of the ICU stay, the readmission task uses clinical information from the index hospitalization preceding discharge. The outcome is defined as a subsequent hospital admission occurring within 30 days after discharge from the index hospitalization.

The similar trajectory construction, feature preprocessing, training/test separation, and L1-regularized logistic regression methodology described for the ICU mortality case were applied. Since the effect of varying the observation-window duration was investigated extensively in the ICU mortality experiments, we do not repeat this analysis here. Instead, the readmission experiment uses a fixed observation window comprising the last 72 hours of the index hospitalization. This period provides a clinically relevant representation of the patient’s condition and treatment course immediately preceding discharge.

#### 4.2.1 Dataset and Prediction Performance

Table 7 demonstrates training and test dataset sizes for the 30-day readmission task after all filtering.

**Table 7:** Training and test dataset sizes for the 30-day readmission task.

| Dataset | Number of Encounters | Number of Features |
| --- | --- | --- |
| Training set | 56,055 | 353 |
| Test set | 13,945 | 353 |
| **Total** | **70,000** | **353** |

To investigate the effect of trajectory representation granularity, we considered two numerical trajectory encoding schemes: a *three-state trajectory representation*, which classifies numerical trajectories as `INCREASING, STABLE`, or `DECREASING`, and a *five-state trajectory representation*, which further distinguishes `SLIGHTLY` and `STRONGLY` increasing or decreasing trajectories. The same predictive model and experimental procedure were used for both representations, allowing their effect on prediction performance and predictive feature selection to be compared directly.

Table 8 shows two sets of metrics for these two representations.

**Table 8:** Prediction performance for three-state and five-state numerical trajectory representations.

| Trajectory Representation | Accuracy | F1-score | AUROC |
| --- | --- | --- | --- |
| Three-state trajectory | 0.6427 | 0.4394 | 0.6875 |
| Five-state trajectory | 0.5991 | 0.4391 | 0.6876 |

Figure 7 shows comparison of top 30 features for these two cases.

**Figure 7:**
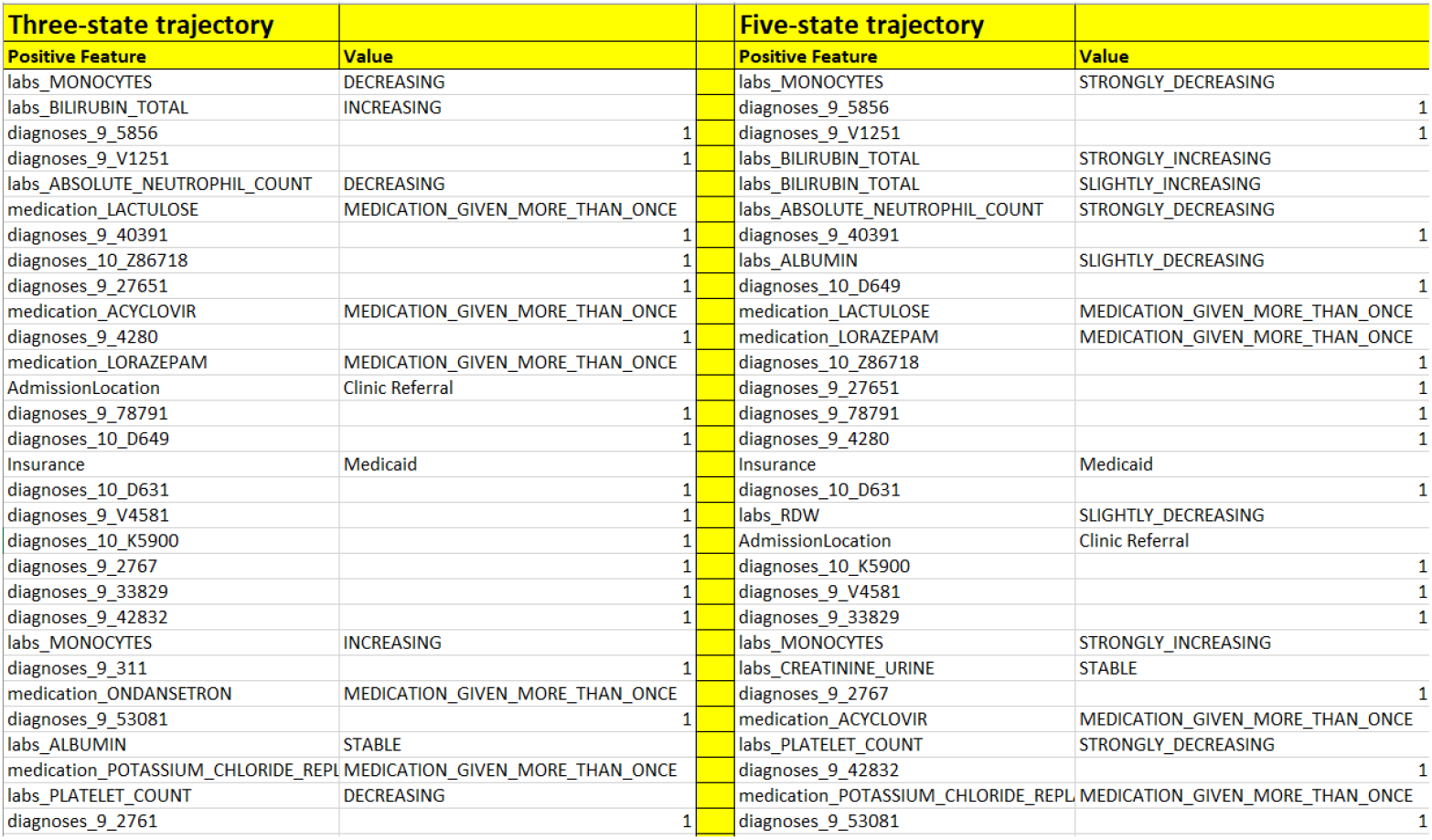
Comparison of top 30 positive numerical features for Three- and Five-state trajectory cases

The comparison between the three-state and five-state numerical trajectory representations shows that increasing the granularity of trajectory classification does not provide a meaningful improvement in predictive performance. The two representations achieve essentially identical AUROC values (0.6875 and 0.6876) and F1-scores (0.4394 and 0.4391), respectively, while the three-state representation achieves higher accuracy (0.6427 versus 0.5991).

Analysis of the selected predictive features leads to a similar conclusion. The two representations identify many of the same clinical features among the strongest predictors, while the five-state representation primarily provides a finer subdivision of numerical trajectories into slightly and strongly increasing or decreasing patterns. In some cases, different trajectory states of the same clinical feature are therefore represented as separate predictors.

These results suggest that the additional granularity of the five-state representation provides limited predictive benefit for the present 30-day readmission task. The simpler three-state representation retains comparable discriminative performance while providing a more compact and directly interpretable description of temporal behavior. Consequently, the three-state representation appears preferable for this study, supporting the use of INCREASING, DECREASING, and `STABLE` as the primary numerical trajectory categories.

## 5 Conclusion

In this work, we proposed an interpretable trajectory-based representation for extracting predictive features from longitudinal structured electronic health record data. Rather than representing temporal information solely through individual measurements, conventional summary statistics, or complex learned representations, the proposed approach transforms the evolution of clinical variables into compact and directly interpretable trajectory states. Numerical measurements are represented by states such as INCREASING, DECREASING, and `STABLE`, while categorical temporal information is represented using clinically meaningful states describing occurrence, repetition, stability, or change. These trajectory features can be combined with static patient and hospitalization characteristics and used directly by conventional predictive models.

The proposed representation was evaluated using MIMIC-IV in two clinically and temporally distinct prediction tasks: in-hospital mortality following ICU admission and hospital readmission within 30 days after discharge. In the ICU mortality study, observation windows ranging from 1 to 48 hours were examined. The experiments demonstrated that temporal trajectory features make a substantial contribution to prediction and that their predictive contribution evolves as a longer portion of the patient’s ICU course becomes available. The best discrimination was obtained with a 32-hour observation window, with an AUROC of 0.8751, while the 24-hour window provided a favorable overall combination of predictive metrics. Analysis across observation windows also identified predictive features that remained consistently highly ranked, providing an additional mechanism for identifying robust and clinically interpretable patterns associated with mortality.

The 30-day readmission study provided a complementary evaluation using clinical information from the final 72 hours of the index hospitalization. We additionally compared three-state and five-state numerical trajectory representations to investigate whether greater trajectory granularity improves prediction. The two representations produced essentially identical AUROC values (0.6875 and 0.6876) and F1-scores (0.4394 and 0.4391), while many of the strongest predictive features were shared between them. These results indicate that subdividing increasing and decreasing trajectories into slightly and strongly changing states provides limited additional predictive benefit for this task. The simpler three-state representation therefore offers an attractive balance between predictive information, compactness, and interpretability.

An important characteristic of the proposed framework is that its primary contribution lies in the representation of longitudinal clinical information rather than in a particular machine-learning algorithm. L1-regularized logistic regression was used in this study because its sparse coefficients provide a transparent mechanism for identifying and ranking predictive features. However, the trajectory representation itself is model-independent and can be incorporated into other statistical and machine-learning approaches.

The trajectory definitions investigated here are intentionally simple and capture only selected aspects of temporal evolution. This simplicity is also a useful property: the resulting features can be readily understood and examined without requiring interpretation of complex latent temporal representations. Future work can extend the framework by incorporating additional information about trajectory magnitude, variability, duration, and more complex temporal patterns, while preserving the principle of direct clinical interpretability. Evaluation on additional clinical outcomes, healthcare institutions, and external datasets will also be important for assessing the generalizability of the identified trajectory patterns.

Overall, the results demonstrate that heterogeneous longitudinal EHR data can be transformed into a compact set of human-interpretable temporal features that retain clinically useful predictive information. Such representations provide a practical bridge between longitudinal clinical data and transparent predictive modeling, with the potential to support interpretable risk assessment, predictive feature discovery, and clinical decision-support applications.

## Data Availability

All data produced in the present study are available upon reasonable request to the authors

https://physionet.org/content/mimiciv/3.0/

